# Children’s Anxiety and Physical Activity during COVID-19 in Relation to Prenatal Exposure to Gestational Diabetes

**DOI:** 10.1101/2020.08.06.20169565

**Authors:** Jasmin M. Alves, Alexandra G. Yunker, Alexis DeFendis, Anny H. Xiang, Kathleen A. Page

## Abstract

**Research goal:** Assess the relationships between anxiety levels, physical activity and in utero exposure to Gestational Diabetes mellitus (GDM) in children age 9 to 15, during the coronavirus disease 2019 (COVID-19) pandemic.

**Methods:** During the COVID-19 pandemic, participants completed phone call or video calls with study personnel where they were asked to report on their physical activity and anxiety levels using the 24-hour physical activity recall and the State-Trait Anxiety Inventory for Children. GDM-exposure was assessed using electronic medical records.

**Results:** Children who reported higher levels of moderate to vigorous physical activity or vigorous physical activity, reported lower anxiety symptoms. Children exposed to GDM in utero reported higher anxiety scores and lower engagement in vigorous physical activity compared to unexposed children. Moreover, the pathway through which children exposed to GDM in utero, reported higher anxiety was partially explained by reduced engagement in vigorous physical activity (75%, p=0.05).

**Conclusions:** Engaging in physical activity during the COVID-19 pandemic may be beneficial for reducing anxiety, particularly among children exposed to GDM in utero, who are at increased risk for adverse psychological outcomes.

## Introduction

The coronavirus disease 2019 (COVID-19) pandemic is an unprecedented stressful time period and is associated with adverse mental health outcomes ^1^. Correspondingly, prenatal exposure to gestational diabetes mellitus (GDM) is associated with increased risk for psychiatric disorders ^2^, and children exposed prenatally to GDM may be particularly vulnerable to heightened anxiety during the pandemic. Moreover, engaging in physical activity (PA) is associated with mental health benefits ^1^, but children are engaging in reduced PA during the pandemic ^3^. Our goal was to investigate children’s anxiety and PA levels during the COVID-19 pandemic in relation to prenatal GDM-exposure.

## Methods

Participants were recruited from the existing observational *BrainChild* study on neuroendocrine programming associated with GDM-exposure. Children were born at a Kaiser Permanente Southern California (KPSC) and had no history of significant medical or psychiatric disorders. Institutional Review Boards (IRB) at the University of Southern California (#HS-15-00540) and KPSC (#10282) approved this study.

Each mother’s GDM status was determined from electronic medical records. The study included one phone/video call visit with both the participant and a parent present between April 20^th^-June 26^th^, 2020. Questionnaires were read aloud by a trained staff member, and the participant gave answers verbally.

Physical activity was assessed using a 24-h physical activity recall (PAR), using metabolic equivalent (MET) values were from the Compendium of Physical Activities ^4^. METs ≥3 were classified as moderate-to-vigorous physical activity (MVPA) and MET ≥6 as vigorous physical activity (VPA).

State anxiety (S-Anxiety) was assessed via the State-Trait Anxiety Inventory for Children (STAIC) ^5^. The S-Anxiety sub-scale was completed to investigate how children were acutely responding to the pandemic.

The relationships between GDM-exposure and S-Anxiety and PA were assessed via linear/logistic regression analysis when appropriate. MVPA was not normally distributed, and square-root transformation was applied. Many children did not engage in VPA (65%), therefore VPA was treated as a categorical variable (yes/no VPA). Maternal education, defined as high school, some college and college/post, was the only covariate significantly different between GDM-exposed and unexposed children, and therefore was included as covariate. In a post-hoc analysis, we examined PA as a mediator for the association between GDM-exposure and S-Anxiety.

## Results

Of the 82 participants from the BrainChild cohort that were eligible to participate, 65 completed the phone call visits. Participant characteristics are described in **Table 1**.

**Table 1.**
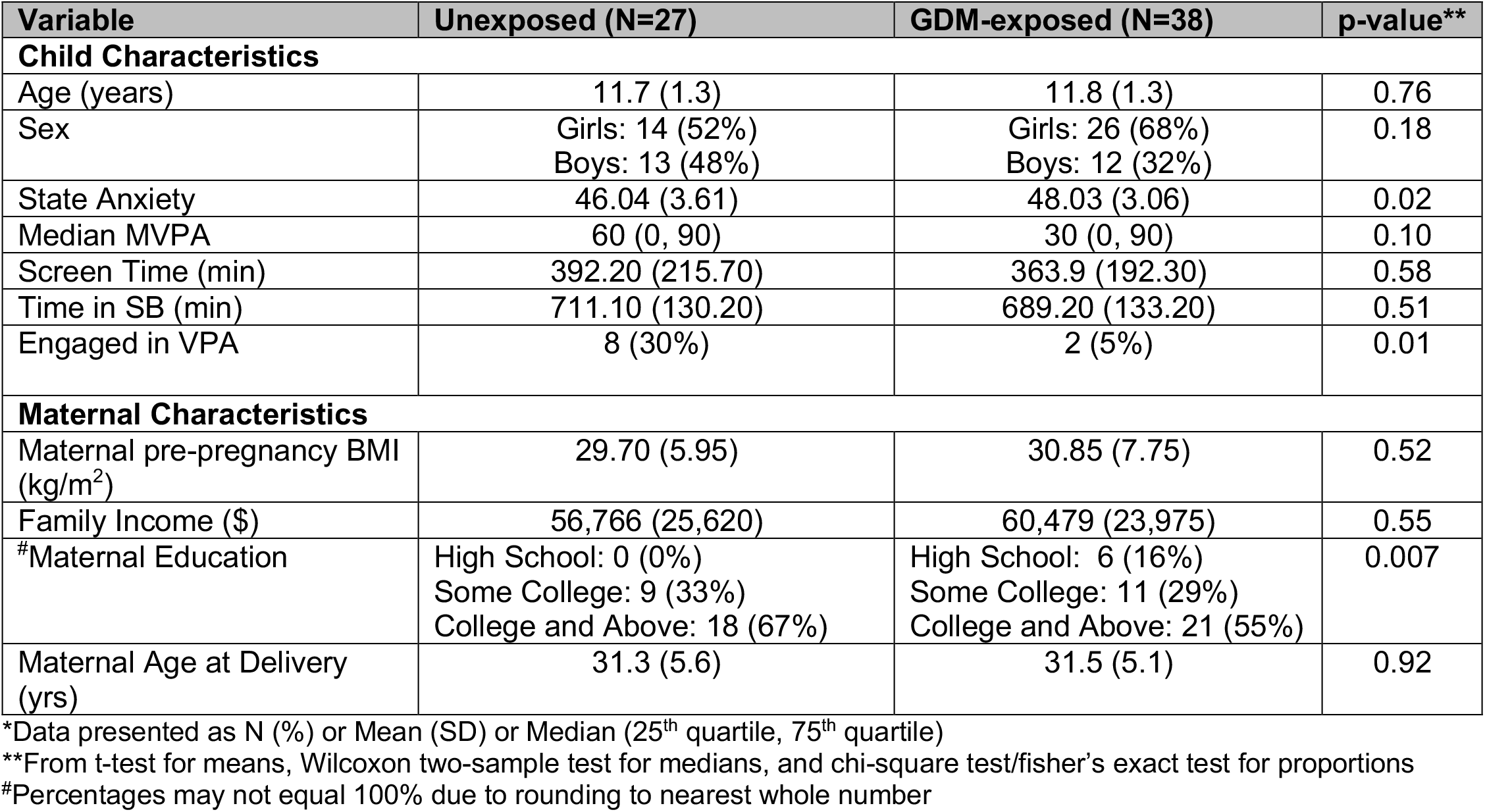
Participant and Maternal Characteristics*.

| Variable | Unexposed (N=27) | GDM-exposed (N=38) | p-value** |
| --- | --- | --- | --- |
| **Child Characteristics** |  |  |  |
| Age (years) | 11.7 (1.3) | 11.8 (1.3) | 0.76 |
| Sex | Girls: 14 (52%)<br>Boys: 13 (48%) | Girls: 26 (68%)<br>Boys: 12 (32%) | 0.18 |
| State Anxiety | 46.04 (3.61) | 48.03 (3.06) | 0.02 |
| Median MVPA | 60 (0, 90) | 30 (0, 90) | 0.10 |
| Screen Time (min) | 392.20 (215.70) | 363.9 (192.30) | 0.58 |
| Time in SB (min) | 711.10 (130.20) | 689.20 (133.20) | 0.51 |
| Engaged in VPA | 8 (30%) | 2 (5%) | 0.01 |
| **Maternal Characteristics** |  |  |  |
| Maternal pre-pregnancy BMI (kg/m<sup>2</sup>) | 29.70 (5.95) | 30.85 (7.75) | 0.52 |
| Family Income (\$) | 56,766 (25,620) | 60,479 (23,975) | 0.55 |
| #Maternal Education | High School: 0 (0%)<br>Some College: 9 (33%)<br>College and Above: 18 (67%) | High School: 6 (16%)<br>Some College: 11 (29%)<br>College and Above: 21 (55%) | 0.007 |
| Maternal Age at Delivery (yrs) | 31.3 (5.6) | 31.5 (5.1) | 0.92 |

\*Data presented as N (%) or Mean (SD) or Median (25<sup>th</sup> quartile, 75<sup>th</sup> quartile)
\*\*From t-test for means, Wilcoxon two-sample test for medians, and chi-square test/fisher's exact test for proportions
#Percentages may not equal 100% due to rounding to nearest whole number

GDM-exposed children reported greater S-Anxiety (regression coefficient *ß*=2.0, p=0.02), compared to unexposed children **(Table 1)**. The proportion of children who engaged in any VPA was significantly lower for GDM-exposed children (5%) compared to unexposed children (30%), (p=0.01). MVPA levels did not differ between GDM-exposed and unexposed children (p=0.10), but GDM-exposed children tended to engage in less MVPA **(Table 1)**. Adjusting for maternal education did not change the results appreciably (p=0.05, 0.03, and 0.07 for S-anxiety, VPA, and MVPA, respectively). Children who reported more time spent in MVPA had lower S-Anxiety (r=-0.34, p<0.01). Children who engaged in VPA also had lower S-Anxiety (mean±SD, 44.8±3.2) compared to children who did not engage in any VPA (mean±SD, 47.6±3.3), (p=0.01). Accounting for MVPA levels explained 19% (p=0.26) of the relationship between GDM-exposure and S-Anxiety, while accounting for VPA levels explained 75% (p=0.05) of the relationship.

## Discussion

During the pandemic, children exposed to GDM reported higher anxiety symptoms and less engagement in VPA compared to unexposed children. Moreover, we found that the pathway between GDM-exposure and greater anxiety in children was partially explained by lower engagement in VPA.

Our findings suggest that engaging in VPA during stressful periods may be beneficial for reducing anxiety levels, particularly among GDM-exposed children, who may be more vulnerable to adverse psychological outcomes. However, larger sample sizes are needed to replicate our findings.

## Data Availability

The datasets generated during and analyzed during the current study are available from Dr. Page the corresponding author, on reasonable request.

## Acknowledgments

The authors would like to thank the families who participate in the BrainChild Study. The authors would also like to thank Ana Romero for managing the BrainChild study, Mayra Martinez and Janet Mora-Marquez for recruiting volunteers and helping collect participant data.

## Author Contributions

Dr. Page and Dr. Xiang had full access to all the data in the study and take responsibility for the integrity of the data and accuracy of the data analysis.

Dr. Page and Dr. Xiang contributed to study concept and design.

Dr. Page, Dr. Xiang, Dr. Alves and Ms. Yunker drafted the manuscript.

All authors critically revised the manuscript for intellectual content.

Ms. Yunker and Ms. Defendis collected and organized data.

Dr. Xiang and Dr. Alves performed the statistical analyses.

Dr. Page and Dr. Xiang obtained funding and provided study supervision.

